# Sex-specific genetic and transcriptomic liability to neuroticism

**DOI:** 10.1101/2022.03.03.22271772

**Authors:** Frank R Wendt, Gita A Pathak, Kritika Singh, Murray B Stein, Karestan C Koenen, John H Krystal, Joel Gelernter, Lea K Davis, Renato Polimanti

## Abstract

**Background:** The presentation, etiology, and relative risk of psychiatric disorders are strongly influenced by biological sex. Neuroticism is a transdiagnostic feature of psychiatric disorders displaying prominent sex differences. We performed genome-wide association studies (GWAS) of neuroticism separately in males and females to identify sex-specific genetic and transcriptomic profiles.

**Methods:** Neuroticism scores were derived from the Eysenck Personality Inventory Neuroticism scale. GWAS were performed in 145,669 females and 129,229 males from the UK Biobank considering autosomal and X-chromosomal variation. Two-sided Z-tests were used to test for sex-specific effects of discovered loci, genetic correlates (N=673 traits), tissue and gene transcriptomic profiles, and polygenic associations across health outcomes in the Vanderbilt University Biobank (BioVu, 39,692 females and 31,268 males).

**Results:** The SNP-heritability of neuroticism was not statistically different between males (*h*^*2*^=10.6%) and females (*h*^*2*^=11.85%). Four female-specific (rs10736549-*CNTN5*, rs6507056-*ASXL3*, rs2087182-*MMS22L*, and rs72995548-*HSPB2*) and two male-specific (rs10507274-*MED13L* and rs7984597) neuroticism risk loci reached genome-wide significance. Male- and female-specific neuroticism polygenic scores were most significantly associated with “mood disorders” (male OR=1.11, *P*=1.40×10^−9^; female OR=1.14, *P*=6.05×10^−22^). They also associated with sex-specific laboratory measures related to erythrocyte count, distribution, and hemoglobin concentration. Gene expression variation in the pituitary was enriched for neuroticism loci in males (males *β*=0.026, *P*=0.002) and genetically-regulated transcriptomic changes highlighted the effect of *RAB7L1, TEX26*, and *PLOT1*.

**Conclusions:** Through a comprehensive assessment of genetic risk for neuroticism and the associated biological processes, this study identified several molecular pathways that can partially explain the known sex differences in neurotic symptoms and their psychiatric comorbidities.

## INTRODUCTION

Neuroticism is a personality trait defined as a tendency to experience negative affect including anger, anxiety, self-consciousness, irritability, emotional instability, and depression in response to perceived threat or punishment (1). Neurotic symptoms are shared among psychiatric diagnoses such as posttraumatic stress disorder (PTSD), generalized anxiety disorder (GAD), and major depressive disorder (MDD). Due to its transdiagnostic characteristics, genome-wide association studies (GWAS) of neuroticism may detect molecular targets relevant across diagnostic categories. Accordingly, genetic factors associated with neuroticism may also be associated with several psychiatric diagnoses.

Biological sex plays a major role in the complexity of this personality trait. In large studies, including those across cultural boundaries (2), women scored higher than men on tests for neuroticism (3,4). These differences were small to moderate depending on the cultural context, but statistical differences persisted regardless of test country (2). This sex difference also extends to the disorders partially overlapping with neurotic symptoms. For example, women score higher than men on indices of GAD, and the lifetime incidence of MDD in women is almost twice that of men (5,6). Women are nearly four times more likely than men to develop PTSD also accounting for differences in traumatic experiences (7).

The twin and SNP-heritability (*h*^*2*^) of neuroticism range from 30-50% (8) and 6-15% (9-12), respectively. Large GWAS conducted in sex-combined cohorts have discovered up to 150 independent SNPs associated with neuroticism (9,11,13-16). These discoveries consistently implicate neuronal genesis and differentiation as well as serotonergic and dopaminergic signaling (14,17). The frontal cortex is frequently identified as a region of interest for neuroticism and related psychopathologies. Furthermore, the worry and anxiety features of neuroticism appears to be dissociable and represent two distinct ends of the neuroticism spectrum (14). In combination, these previous studies inform the epidemiologic differences in neuroticism and provide clues to understanding individual liability to poor mental health as it relates to negative affect. Despite the known phenotypic and epidemiologic difference by sex, it remains unclear if the genetic vulnerability to neuroticism differs across sexes.

Genetic studies of sex differences in neuroticism have been performed on the X-chromosome. Seven genes on the X-chromosome were previously associated with neuroticism including *HS6ST2* (13). X-chromosome variants accounted for 0.22-0.43% of the variance in neuroticism depending on the method used to adjust for dose of the X-chromosome by sex (13). Furthermore, sex-stratified X-chromosomal polygenic scores were unable to predict neuroticism highlighting that autosomal chromosomes account for the majority of the sex differences in the heritability of this personality trait.

Here, we present the first large-scale neuroticism GWAS investigating the genetic component of sex differences. This study describes sex-specific risk loci for neuroticism across the genome. These loci contribute to sex-specific transcriptomic and biomarker profiles relevant for disease etiology involving neurotic symptoms.

## METHODS

### Discovery Sample and Neuroticism Definition

The UKB is a population-based cohort of >502,000 participants between 37 and 73 years old at the time of recruitment. UKB assesses a wide range of factors including physical health, anthropometric measurements, circulating biomarkers, and sociodemographic characteristics. The UKB consists of 273,366 female and 229,114 male participants classified into six ancestry groups.

The neuroticism score was derived from 12 neurotic behavior questions from the Eysenck Personality Inventory Neuroticism scale (18,19). At the time of their baseline survey, participants were asked to answer each question with “*yes*,” “*no*,” “*do not know*,” or “*prefer not to answer*.” Neuroticism questions were: “*Does your mood often go up and down*?”, “*Do you ever feel ‘just miserable’ for no reason*?”, “*Are you an irritable person*?”, “*Are your feelings easily hurt*?”, “*Do you often feel ‘fed-up’*?”, “*Would you call yourself a nervous person*?”, “*Are you a worrier*?”, “*Would you call yourself tense or ‘highly strung’*?”, “*Do you worry too long after an embarrassing experience*?”, “*Do you suffer from ‘nerves’*?”, “*Do you often feel lonely*?”, “*Are you often troubled by feelings of guilt*?”. The neuroticism score is the sum of all “yes” responses per participant and ranges from 0-12 with 12 being the most severe (UKB Field ID 20127).

### Genome-wide Association with Neuroticism

Genotyping and imputation of the UKB have been previously described (20). Briefly, UKB participants were genotyped using a custom Axiom array capturing genome-wide genetic variation and short insertion/deletions, including coding variants and markers providing good coverage for imputation in European ancestry populations. UKB was imputed using the Haplotype Reference Consortium reference panel.

Our GWAS were restricted to unrelated participants of European ancestry as identified by a two-stage ancestry assignment and pruning procedure from the Pan-UKB (details available at https://pan.ukbb.broadinstitute.org/). Linear regression was performed in PLINK-2.0 (21) using SNPs with imputation INFO scores >0.8, minor allele frequencies >0.01, missingness <0.05, and Hardy-Weinberg equilibrium *P*-values >1×10^−10^. We included age and the first 15 within-ancestry principal components as covariates. After quality control, we analyzed genetic data from 145,669 females and 129,229 males.

X-chromosome association tests were performed using XWAS v3.0 (22). X-chromosome SNPs were included in the analysis if they had minor allele frequencies >0.05 and were not significantly different in males and females (*P*<0.05) or missingness <0.05 per sex. Sex-stratified genetic association analysis was performed using PLINK X-chromosome model 2 which codes male X-chromosomal genotypes as 0/2 corresponding to the female homozygote state (23).

### Cross-Ancestry Polygenic Scoring

PRSice-v2 (24) was used to test the cross-ancestry portability of polygenic scores (PGS) derived from male- and female-stratified GWAS of neuroticism. Linkage disequilibrium-independent SNPs were ascertained based on the 1000 Genomes Project (1kGP) European reference using *r*^*2*^=0.01 in 250-kb windows. Regression models were covaried with age and the first ten principal components of ancestry in the target population. Ten *P*-value thresholds (*P*_⊤_) were tested: 1, 0.5, 0.3, 0.1, 0.05, 0.005, 5×10^−4^, 1×10^−5^, 5×10^−6^, 5×10^−7^, and 5×10^−8^. Five populations from the Pan-UKB analysis (details available at https://pan.ukbb.broadinstitute.org/) were tested: African (1,757 males and 2,505 females), Admixed American (252 males and 459 females), Central/South Asian (2,704 males and 2,283 females), East Asian (573 males and 1,087 females), and Middle Eastern (537 males and 406 females).

### Linkage Disequilibrium Score Regression (LDSC)

Observed-scale heritability (*h*^*2*^) was calculated for each GWAS using LDSC (25) with the 1kGP European ancestry reference LD panel. LDSC also was used to calculate the genetic correlation (*r*_*g*_) between (i) male and female neuroticism score GWAS and (ii) between male or female neuroticism GWAS and 673 sex-stratified summary statistics from the UKB (details available at https://github.com/Nealelab/UK_Biobank_GWAS) with significant SNP-*h*^*2*^ in either sex. We excluded male- and female-specific traits so that each *r*_*g*_ could be assessed across sex. Multiple testing correction was applied using the false discovery method (FDR<5%) considering all trait pairs tested: 2 sexes x 673 phenotypes=1,346 total *r*_*g*_ tested.

To test which functional categories contribute to the *h*^*2*^ of neuroticism, we used LDSC to partition each *h*^*2*^ estimate using 118 genomic annotations. All genomic annotations for partitioned *h*^*2*^ have been described previously (26-28) and in the Supplementary Material. Because of the nature of the test, p-values derived from partitioned *h*^*2*^ multiple regressions were not subjected to multiple testing correction (26).

### Functional Annotation of Risk Loci

Neuroticism liability loci were positionally mapped with Multi-marker Analysis of GenoMic Annotation (MAGMA v1.08) implemented in FUMA v1.6a (29) using 2-kb positional mapping around each lead SNP (30). Linkage disequilibrium independent genomic risk loci are defined by their lead SNP (*P*<5×10^−8^) and all surrounding SNPs with *r*^*2*^>0.6 with the lead SNP. Enrichment of tissue transcriptomic profiles was tested relative to GTEx v8. Multiple testing correction was applied using FDR (5%) considering all tissue-types or cell-types tested in both sexes.

### PGS in BioVu

Phenome-wide (PheWAS) and lab-wide association studies (LabWAS) of neuroticism polygenic scores (PGS) were performed in a sex-stratified manner. Neuroticism polygenic scores with continuous shrinkage were calculated for BioVU participants using PRS-CS (31). PRS-CS is a Bayesian polygenic prediction method that imposes continuous shrinkage priors on SNP effect sizes. LD-independent SNPs were selected based on the 1kGP European ancestry reference panel (31,32).

Female- and male-specific PGS were associated with diagnoses, symptoms, and lab measurements measured on European ancestry patients treated at the Vanderbilt University Medical Center. Phenotypes were extracted from electronic health records (EHRs) and genetic data was available within a biobank lined to the HER called BioVU. Phenotypes in the EHR are represented as phecodes, which represent binary hierarchical clustering of the International Classification of Diseases (ICD9/ICD10) codes (33,34). A minimum number of 100 cases were required for inclusion in our PheWAS and a minimum of 100 individuals for inclusion in LabWAS. We then tested 1,020 phecodes within the male-specific strata up to 31,785 individuals) and 1,136 phecodes within the female-specific strata (up to 40,417 individuals). We required a minimum of 100 individuals with a measurement that passed quality control (32) for a given laboratory test to be included in the LabWAS. We then tested 299 labs in the male-stratified analysis (up to 31,268 individuals) and 303 labs in the female-stratified analysis (up to 39,692 individuals). We employed the PheWAS package in R (see https://github.com/PheWAS/PheWAS) to perform linear or logistic regression to identify the labs and phecodes that were associated with neuroticism PGS after adjusting for age and the top ten principal components from genetic data to control for population stratification. For LabWAS and PheWAS, multiple testing correction was applied by sex using FDR (5%) to account for the correlation among phecodes.

### Transcriptome-wide Association Study (TWAS) in Frontal Cortex and Pituitary

Summary-based TWAS of neuroticism was performed with FUSION (35). FUSION predicts the relationship between gene expression and a trait using a linear combination of GWAS Z-statistics and a set of functional weights derived from GTEx v7 prefrontal cortex and pituitary. Weights for 3,144 features in the prefrontal cortex were based on 118 samples and weights for 4,402 features in the pituitary were based on 157 samples. For each gene, weights are derived from one of five models that produces the largest *R*^*2*^ between predicted and observed expression. Multiple testing correction was applied using a stringent Bonferroni threshold (*P*<1.59×10^−5^=0.05/3,144 genes for prefrontal cortex and *P*<1.14×10^−5^=0.05/4,402 genes for pituitary).

### Gene Expression Effects across Developmental Stage

To further explore TWAS results in the context of sex-differential gene expression, we used the BrainSpan Atlas of the Developing Human Brain (36). BrainSpan consists of postmortem human brain tissue and fetal specimens. Samples were excluded if they had (i) evidence of karyogram, microscopic neuronal, or physical malformations, (ii) maternal substance use during pregnancy, (iii) positive for Hepatitis B, Hepatitis C, and HIV, and (iv) postnatal evidence of excessive maternal substance use, neurological or psychiatric disorder, or multiple organ failure (36).

We used transformed expression values (reads per kilobase per million (RPKM)) (37) from the frontal cortex and calculated the average expression for all TWAS genes as a function of sex, developmental stage, and region:

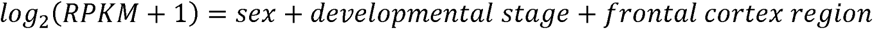

We combined expression data from five frontal cortex regions: anterior (rostral) cingulate (medial prefrontal) cortex, dorsolateral prefrontal cortex, orbital frontal cortex, ventrolateral prefrontal cortex, and the primary motor cortex (36). This tissue selection resulted in 159 frontal cortex tissue samples (N=68 females).

### Statistical Comparisons Across Sex

We evaluated the sex-specificity of a result by comparing the effect estimates (*e*.*g*., SNP-*h*^*2*^, SNP *β*, PGS *β*, or *r*_*g*_) using z-tests for all estimates significant in one sex (FDR<5%) and not nominally significant in the other sex (*P*>0.05):

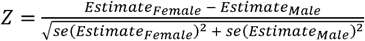

For example, sex-specific risk loci were defined as any locus meeting genome-wide significance threshold (*P*<5×10^−8^) in one sex but failing to reach a nominal significance threshold (*P*<0.05) in the other sex. The effect sizes of these loci also were significantly different between sexes. This procedure was applied to SNP effect sizes, *h*^*2*^, *r*_*g*_, and BioVU PheWAS and LabWAS effect estimates. There were no instances of qualitative heterogeneity in analysis results whereby an association survived multiple testing correction in one sex and had a nominally significant (*P*<0.05) discordant effect in the other sex.

## RESULTS

### Sex-stratified GWAS of neuroticism in autosomes and X-chromosome

The mean neuroticism score for females (4.51±0.008) was greater than males (3.77±0.007; Cohen’s *d*=1.33; Wilcox *P*<2.2×10^−16^; Fig S1).

There were 9 and 14 genome-wide significant (GWS; *P*<5×10^−8^) LD-independent loci identified in male and female GWAS of neuroticism, respectively (Fig 1a, Fig S2, and Tables S1 and S2). The most significantly associated locus in both sexes (males rs62062288, *P*=6.06×10^−13^; females rs55955207, *P*=3.62×10^−12^) positionally mapped to a gene-dense region of chromosome 17 containing targets previously implicated in other disorders and internalizing psychopathologies such as *CRHR1, ARHGAP27, PLEKHM1*, and *MAPT* (among others).

**Fig 1.**
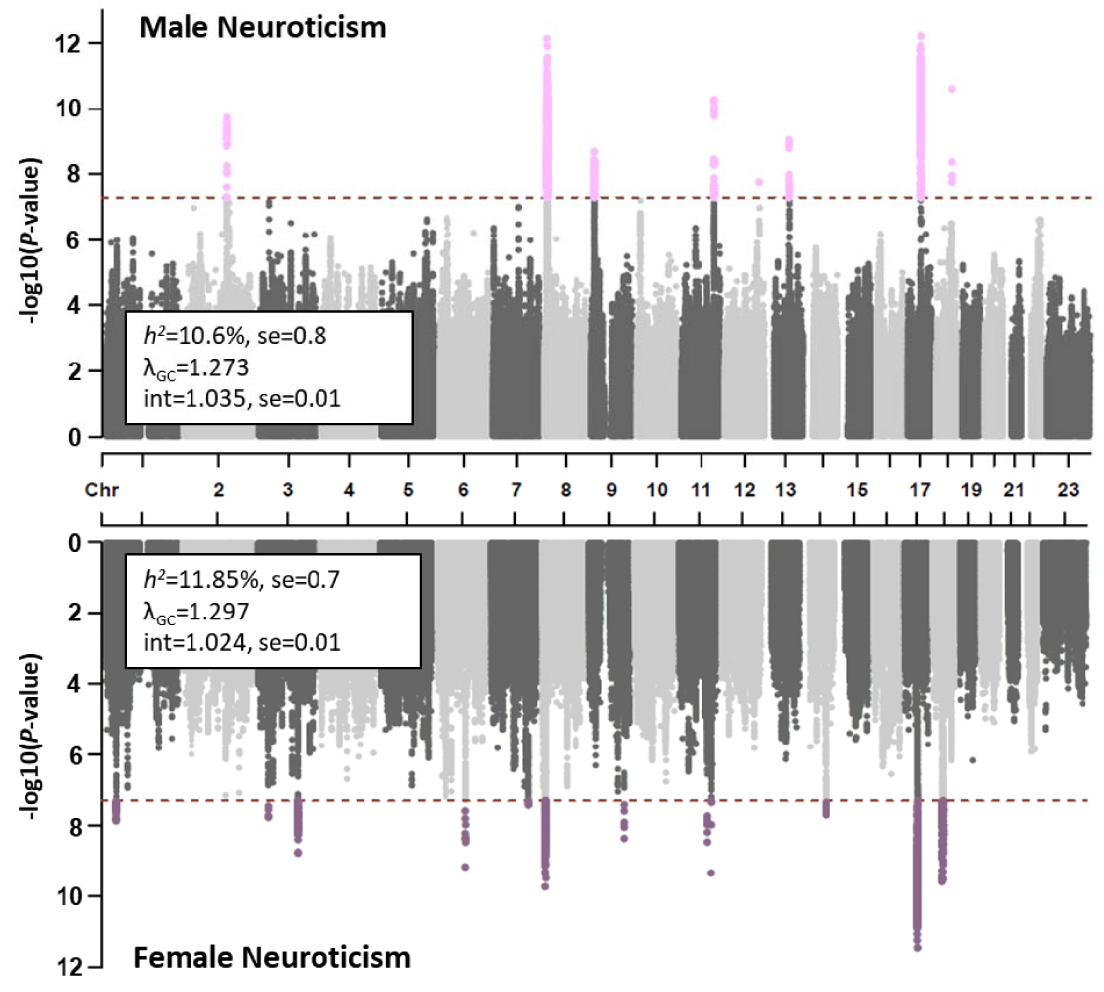
Sex-stratified GWAS and locus differences. Manhattan plots for neuroticism in males (top) and females (bottom) with GWAS quality statistics embedded in each. Sex specificity of identified risk loci are shown in Table 1.

Of the 23 loci associated with neuroticism, six met criteria for sex-specific effects as they (i) met GWS in one sex and were not associated with neuroticism in the other sex (*P* > 0.05) and (ii) had significant differences in effect size in males and females (Fig 1b and Table 1). The two male-specific loci were rs10507274*T (*MED13L*-*C12orf49*-*RNFT2* gene cluster) and rs7984597*A (intergenic), both of which associated with lower neuroticism scores. The four female-specific loci were rs10736549*A (*CNTN5*), rs2087182*G (*MMS22L*), rs6507056*G (*ASXL3*-*NOL4* gene cluster), and rs72995548*C (*HSPB2*-*PIHID2*-*CBO2*-*PTS* gene cluster). There was no evidence of qualitative heterogeneity (i.e., opposite effect directions) among neuroticism risk loci. We did not identify X-chromosomal loci associated with neuroticism by either genome-wide or chromosome-specific multiple testing correction.

**Table 1.** Sex-specific risk loci for neuroticism in UK Biobank participants of European ancestry. Differences in effect size estimates between sexes were calculated using two-sided Z-tests (see Methods) for each locus (Table S1) meeting genome-wide significance (P < 5×10^−8^) in one sex and failing to reach nominal significance in the other sex.

| Specificity | SNP | Effect Allele | Gene * | Females | | Males | | $P_{\text{sex-diff}}$ |
| --- | --- | --- | --- | --- | --- | --- | --- | --- |
| | | | | $\beta(\text{se})$ | $P$ | $\beta(\text{se})$ | $P$ | |
| Female | rs10736549 | A | *CNTN5* | -0.072 (0.012) | $3.52 \times 10^{-9}$ | -0.002 (0.013) | 0.887 | $6.85 \times 10^{-5}$ |
| | rs6507056 | G | *ASXL3* | -0.077 (0.012) | $2.51 \times 10^{-10}$ | -0.010 (0.013) | 0.411 | $1.51 \times 10^{-4}$ |
| | rs2087182 | G | *MMS22L* | -0.112 (0.018) | $6.76 \times 10^{-10}$ | -0.020 (0.019) | 0.301 | $4.11 \times 10^{-4}$ |
| | rs72995548 | C | *HSPB2* | 0.150 (0.024) | $4.61 \times 10^{-10}$ | 0.040 (0.025) | 0.113 | 0.002 |
| Male | rs10507274 | T | *MED13L* | -0.026 (0.025) | 0.295 | -0.145 (0.026) | $1.68 \times 10^{-8}$ | $8.03 \times 10^{-4}$ |
| | rs7984597 | A | (intergenic) | -0.023 (0.012) | 0.060 | -0.077 (0.013) | $8.88 \times 10^{-10}$ | 0.002 |

\* Comprehensive SNP positional-mapping results are provided in Table S2.

The *h*^*2*^ of neuroticism was 10.6%±0.8 in males and 11.9%±0.7 in females (*P*_diff_=0.220; Fig 1a). The *r*_*g*_ between them supports substantial overlap of the polygenic signal: *r*_*g*_ =0.906, *P*=2.40×10^−278^.

### Cross-Ancestry Polygenic Scoring

In both males and females, the neuroticism PGS derived from a European-descent sample associated with neuroticism in Central/South Asians across almost all thresholds (male N=2,704, *R*^*2*^=0.785%, *P*=3.46×10^−6^, P_⊤_=0.3; female N=2,283, *R*^*2*^=0.709%, *P*=4.87×10^−5^, P_⊤_=0.5; Table S3) with no differences in the effect estimates. Conversely, in the admixed American samples, neuroticism PGS was associated with neuroticism among females (N=459; *R*^*2*^=1.60%, *P*=0.007) but not males (male N=252, *R*^*2*^=0.551%, *P*=0.247; *P*_diff_=0.005) in line with the different sample size available in these two groups. No PGS association survived multiple testing correction in the other ancestry groups tested.

### Partitioned Heritability

Neuroticism SNP-*h*^*2*^ was partitioned using 119 genomic annotations. Male and female SNP-*h*^*2*^ were enriched for 10 and 22 genomic annotations (FDR<0.05), respectively. The most significantly enriched annotation from both GWASs was the Genomic Evolutionary Rate Profiling (GERP) neutral rate score (male enrichment=1.88, *P*=5.45×10^−12^; female enrichment=1.85, *P*=4.15×10^−18^). As the estimated enrichments in males and females were similar (weighted slope=0.982, *P*=1.52×10^−23^, Fig S3), there were no significant differences in enrichment estimates (Table S4).

### Genetic Correlation

We tested the *r*_*g*_ between neuroticism and 673 traits from UKB with *h*^*2*^ Z-scores>4 in either sex (Table S5). The GWASs of neuroticism in males and females were genetically correlated with 172 and 330 traits, respectively (FDR<0.05). In addition to the 12 items contributing to the neuroticism score derivation, the greatest magnitude *r*_*g*_ for males was “*substances taken for depression*” (UKB Field ID 20546_3, *r*_*g*_=0.804, *P*=1.25×10^−16^) and for females was “*substances taken for anxiety*” (UKB field ID 20549_3, *r*_*g*_ =0.812, *P*=8.65×10^−18^). There were no significant differences in *r*_*g*_ estimates between males and females as the magnitude of these estimates strongly overlapped (weighted slope=0.986, *P*=8.15×10^−234^; Fig S4).

### PGS associations in BioVU

To contextualize our findings with broader clinical consequences, neuroticism PGS derived from sex-stratified GWAS were tested for association with 942 phecodes and 290 lab measures from BioVU. Male- and female-specific neuroticism PGS were most significantly associated with “mood disorders” (male OR=1.11, *P*=1.40×10^−9^; female OR=1.14, *P*=6.05×10^−22^; Fig 2 and Table S6). There were no sex-specific findings with respect to BioVU phecodes.

**Fig 2.**
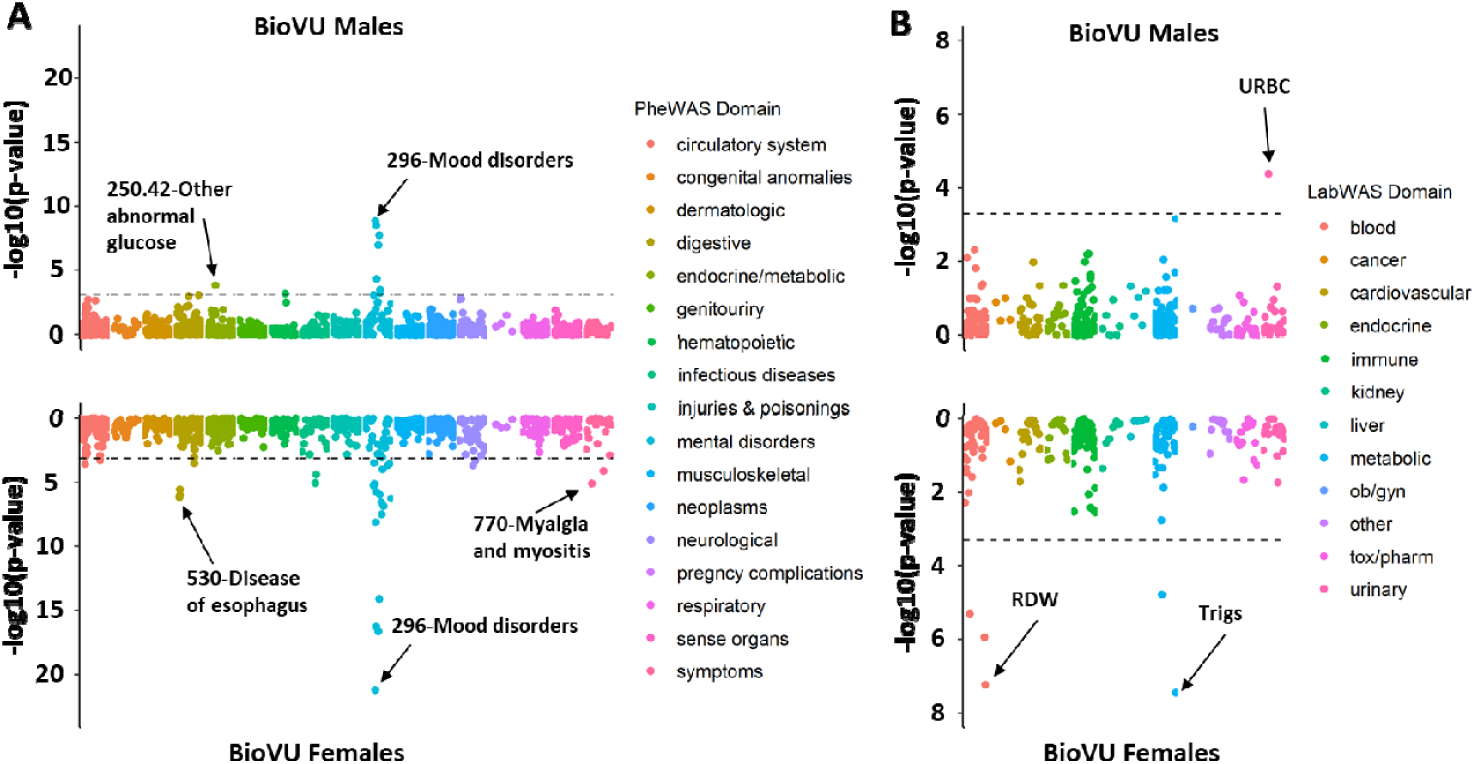
PheWAS and LabWAS results. Manhattan plots of sex-stratified phenome-wide (PheWAS in panel A) and laboratory-wide association studies (LabWAS in panel B) of neuroticism polygenic scores in the Vanderbilt University Biobank (BioVU). Each data point represents a single phecode (PheWAS) or laboratory measurement (LabWAS) with sample sizes per data point provided in Tables S6 and S7. Abbreviations: RDW=red blood cell distribution width; Trigs: triglyceride concentration; URBC=urinary red blood cell count.

The most significant laboratory measurements associated with neuroticism were erythrocytes in the urine for male PGS (*β*=-0.035, *P*=4.15×10^−5^) and triglyceride concentration for female PGS (*β*=0.040, *P*=3.58×10^−8^). The BioVU LabWAS highlighted several sex-differentiated lab measures (Table 2 and Table S7).

**Table 2.** Sex-specific associations between neuroticism PGS in males and females and laboratory measures from Vanderbilt BioVU.

| Specificity | BioVU Laboratory Measurement | Female | | | Male | | | $P_{\text{sex-diff}}$ |
| --- | --- | --- | --- | --- | --- | --- | --- | --- |
| | | $\beta(\text{se})$ | p | N | $\beta(\text{se})$ | p | N | |
| Male | Erythrocytes [# / area] in urine sediment by microscopy high power field | -0.006 (0.007) | 0.382 | 14764 | -0.035 (0.008) | $4.15 \times 10^{-5}$ | 10628 | 0.012 |
| Female | Mean corpuscular hemoglobin [mass/volume] by automated count | -0.021 (0.005) | $4.91 \times 10^{-6}$ | 33474 | 0.010 (0.006) | 0.102 | 26110 | $3.89 \times 10^{-5}$ |
| | Erythrocyte distribution width [entitic volume] by automated count | 0.024 (0.005) | $1.17 \times 10^{-6}$ | 35100 | -0.007 (0.006) | 0.228 | 28142 | $5.55 \times 10^{-5}$ |
| | Erythrocyte distribution width [Ratio] by Automated count | 0.026 (0.005) | $5.96 \times 10^{-8}$ | 39692 | 0.003 (0.006) | 0.617 | 31267 | 0.002 |
| | Calcium serum/plasma serum/plasma | -0.019 (0.005) | $1.69 \times 10^{-5}$ | 37389 | 0.004 (0.005) | 0.410 | 30527 | $4.54 \times 10^{-4}$ |

### Frontal Cortex and Pituitary Transcriptomic Profile Enrichment

The most significant transcriptomic profiles enriched in GWAS of neuroticism was frontal cortex (BA9; males *β*=0.025, *P*=1.93×10^−4^; females *β*=0.028, *P*=5.63×10^−5^) and cerebellum (males *β*=0.015, *P*=0.010; females *β*=0.029, *P*=6.61×10^−6^), respectively (Table S8). There was strong correlation between the male and female frontal cortex TWAS (slope=0.951, *P*=1.03×10^−122^; Fig 3A and Table S9).

**Fig 3.**
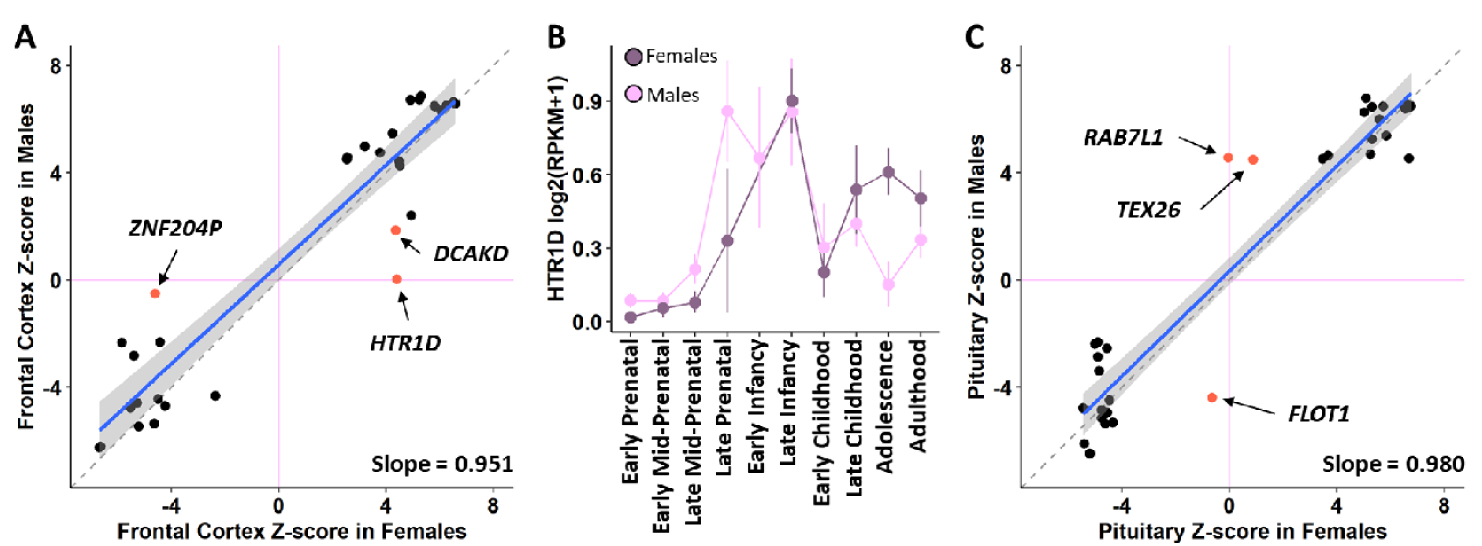
TWAS comparison. (A) Relationship between male and female frontal cortex TWAS Z-scores for all significant associations after multiple testing correction in either sex (*P* < 1.59×10^−5^). Each data point is a single gene associated with neuroticism in either males or females. Purple lines indicate z-score of 0 (no association). The dashed grey line denotes the 1-to-1 relationship between males and females and the blue line shows the linear regression between sexes. Labeled genes (in red) are significantly associated with neuroticism in one sex but do not meet nominal significance in the other. (B) Sex-by-developmental stage effects on frontal cortex gene expression (error bars denote 95% confidence interval) from BrainSpan (36). Sample sizes per developmental stage are provided in Table S10. (C) Relationship between male and female frontal cortex TWAS Z-scores for all significant associations after multiple testing correction in either sex (*P* < 1.14×10^−5^).

Three frontal cortex TWAS genes were associated with neuroticism in females but did not reach nominal significance in males: *ZHN204P, HTR1D*, and *DCAKD* (Table S10). To evaluate the sex-differential expression of each gene, we compared male and female mean expression values from BrainSpan (36). Sex was independently associated with *HTR1D* expression in the frontal cortex (*β*=-0.194, *P*=0.021) but also exhibited an interaction effect between sex and developmental stage (*β*=0.036, *P*=0.007; Fig 3B).

There was a male-specific enrichment of pituitary transcriptomic profiles (males *β* =0.026, *P*=0.002; females *β*=5.48×10^−4^, *P*=0.477, *P*_diff_=0.003). To identify genes underlying this sex-differential enrichment, we tested the predicted expression of 4,402 genes in the pituitary for association with neuroticism. There was a strong correlation between the pituitary TWAS from males and females (slope=0.980, *P*=3.99×10^−93^; Fig 3C and Table S11) and most overlapping TWAS signal reflects shared effects of the chromosome 17 *CRHR1*- and *WNT3*-*KANSL1*-*ARL17A*-containing loci often implicated in disorders with internalizing symptoms (38,39). Three genes detected in males (*P*<1.14×10^−5^) were not identified in females (*P*>0.05): *RAB7L1* (males: *Z*=4.56, *P*=5.00×10^−6^; females *Z*=-0.045, *P*=0.964), *FLOT1* (males: *Z*=-4.39, *P*=1.11×10^−5^; females *Z*=-0.639, *P*=0.523), and *TEX26* (males: *Z*=4.48, *P*=7.46×10^−6^; females *Z*=0.986, *P*=0.267).

## DISCUSSION

Although biological sex plays a major role in the presentation of psychiatric symptoms, this variable is often included only as a covariate in many large GWAS. This procedure hindered the discovery of risk loci and associated biological processes underlying the well-established differences in neuroticism and prevalence of some mental disorders between males and females. Through GWAS and comprehensive *in silico* analysis, we demonstrated that the genetics of neuroticism largely overlap between males and females. However, we highlighted several exceptions that may be useful to understand the biology underlying the sex difference for this trait, and eventually develop sex-specific therapies. Our findings extend to biological processes that may partially explain sex differences in the presentation, etiology, and/or relative risk or psychiatric disorders with prominent neurotic symptoms (40), including PTSD, MDD, and GAD.

Six loci met our strict definition of sex-specificity such that they had genome-wide significant effects on neuroticism in one sex but not even nominal association in the other (*P*>0.05) and had significantly different effect sizes for the two sexes. One male-specific SNP mapped to *MED13L*, a locus harboring rare *de novo* mutations associated with intellectual disability and psychopathology (41). Two female-specific loci mapped to *CNTN5* and *ASXL3. CNTN5* has been associated with suicidal thoughts and behaviors (42,43) and autism spectrum disorder (ASD) (44). This locus has garnered considerable attention because its GWAS effect translates into model organisms of ASD (44).

We found no risk loci associated with neuroticism on the X-chromosome. Prior studies have identified X-chromosomal neuroticism risk loci that survive sex-based heterogeneity in the GWAS sample (13). Because of X-chromosomal inheritance patterns (i.e., female X-inactivation and male hemizygosity), loci on this chromosome that associate with an outcome may confer biologically relevant effects (13). Our lack of significant associations on the X-chromosome suggests that X-chromosomal loci from previous investigations confer a more global effect on neuroticism with no heterogeneity across sex. The lack of X-chromosome association may suggest that sex differences in neuroticism arise because of neurodevelopmental and/or neuroplastic effects of sex steroids (45). As a transdiagnostic feature of disorders with internalizing symptoms, we might hypothesize that X-chromosomal susceptibility loci for PTSD, MDD, and GAD may have similar global effects with few sex-specific effects (46). This question is a major focus of ongoing consortium efforts of study genetic susceptibility to psychiatric disorders.

In both sexes, neuroticism PGS from European ancestry participants translated to Central/South Asian UKB participants. However; PGS poorly translated to African, East Asian, Admixed American, and Middle Eastern populations (47), highlighting a need for intentional design of mental health studies and participant recruitment from these populations (48).

We also tested for genetic overlap in a phenome-wide manner using two approaches. As expected, we identified strong positive genetic overlap between neuroticism and mood disorders among males and females but no differences could be detected by sex. Using a PGS approach that aggregates neuroticism genetic architecture, we identified sex differences in laboratory measures related to urinary red blood cell count (male-specific) and red blood cell physical properties (e.g., distribution width, mean corpuscular hemoglobin, etc.; female-specific). In males, urinary blood strongly implies kidney or urinary infection and may be an indicator of prostate health (49). The protective effect of neuroticism PGS on urinary blood is opposite to literature of a positive relationship between prostate health and neurotic behavior (50), suggesting the presence of important clinical and environmental factors rather than strong genetic mediation between these traits.

In females, neuroticism PGS was associated with lower mean corpuscular volume and lower red blood cell distribution width, two powerful biomarkers of anemia (51,52). The relationship between anemia and mental health is strong with studies across the lifespan correlating anemia and greater depressive symptoms (53) and cognitive impairment (54). A study of >50,000 individuals demonstrated that iron-deficient anemias associated with an adjusted hazard ratio of 1.49 and 1.47 for depressive and anxiety disorders, respectively. Regardless of iron supplementation, the female participants had a substantially higher incidence of a psychiatric diagnosis compared to male participants (55). Therefore higher neuroticism PGS in women may prevent common treatment efficacy and exacerbate symptoms such as fatigue (56) and depression (57).

Finally, we investigated transcriptomic profiles of neuroticism in males and females. Consistent with prior TWAS of internalizing disorders and their symptoms, loci in the neuroticism GWAS from males and females had a similar magnitude of association with gene expression in the frontal cortex (58,59). This finding reinforces the effects of executive functioning deficits, cognitive flexibility, and prospective memory in depression and anxiety symptoms. The *HTR1D* gene exhibited a significant sex×developmental stage effect. *HTR1D* encodes the 5-hydroxytryptamine receptor-1D which plays an essential role in serotonin signaling and has been suggested as a possible therapeutic target for depression, though prior studies could not narrow down where in the brain this therapeutic effect may be most relevant (60). We demonstrate that in adult frontal cortex, *HTR1D* is more highly expressed in females than males suggesting HTR1D antagonists may be most effective in female patients.

Enrichment of the pituitary transcriptomic profile was male-specific. We further evaluated this finding using TWAS in pituitary tissue from GTEx and identified three loci with significant effects in males but no effect in females (*P*>0.05): *RAB7L1, TEX26*, and *FLOT1*. RAB7L1 is involved in axon (61) morphology and has been previously associated with schizophrenia, bipolar disorder, neurodegenerative disorders, and another transdiagnostic trait: risk-taking (62,63). *TEX26* encodes the protein testis expressed 26 and was recently associated with obsessive-compulsive disorder (64) in a large international meta-analysis. *FLOT1* was upregulated in brains and peripheral blood of major depressive disorder cases (65) and modulates chronic corticosterone response (66) suggesting it may play a role in disease etiology. In our study of the pituitary, *FLOT1* was significantly down-regulated suggesting potential inverse predictive power of disease etiology. The link between the gene expression in the pituitary and neuroticism may emphasizes the well documented relationships between cortisol released from the hypothalamic– pituitary–adrenal axis, mood, and mental health (67,68), especially as they relate to hormone differences between men and women (69).

Though comprehensively assessing the sex-specificity of neuroticism risk loci and associated biological information, this study has several limitations to consider. First, the GTEx expression weights are the result of models including age and sex as covariates. Therefore, the estimates reported here are those which survive sex-heterogeneity in the GTEx project. Though we were able to follow up brain results in a sex-stratified manner using BrainSpan, future studies will require additional samples to support statistically powerful gene expression studies stratified by sex. Second, sex-stratified studies inherently reduce statistical power of a GWAS by separating the total sample. This reduction in sample size may be prohibitive for GWAS of historically excluded populations. The findings from this study further necessitate dedicated recruitment of women and minorities in studies of mental health (48).

Despite these limitations, we report sex-specific genetic and transcriptomic liability to neuroticism across multiple complementary analyses. These findings, and the associated sex-specific biomarkers detected here, launch multiple hypotheses for testing and understanding the sex differences in presentation, etiology, and relative risk of psychiatric disorders with debilitating neurotic symptoms.

## Data Availability

All data supporting the conclusions of this study are provided as supplementary material. This research was conducted using the UK Biobank Resource (application reference no. 58146). UKB data can be accessed by bona fide researchers through the UKB data access portal.

## ACKNOWLEDGEMENTS

This research was conducted using the UK Biobank Resource (application reference no. 58146). The authors thank the research participants and employees of the UK Biobank for making this work possible. This study was supported by the National Institutes of Health (R21 DC018098, R33 DA047527, and F32 MH122058) and One Mind.

## DISCLOSURES

Drs. Gelernter is named as an inventor on PCT patent application #15/878,640 entitled: “Genotype-guided dosing of opioid agonists,” filed January 24, 2018. Dr. Stein is paid for his editorial work on the journals Biological Psychiatry and Depression and Anxiety, and the health professional reference Up-To-Date; he has also in the past 3 years received consulting income from Actelion, Acadia Pharmaceuticals, Aptinyx, atai Life Sciences, Boehringer Ingelheim, Bionomics, BioXcel Therapeutics, Eisai, Clexio, EmpowerPharm, Engrail Therapeutics, GW Pharmaceuticals, Janssen, Jazz Pharmaceuticals, and Roche/Genentech. Drs. Polimanti and Gelernter are paid for their editorial work on the journal Complex Psychiatry. The other authors have no competing interests to report.

Dr. Krystal’s financial disclosures are as follows:

### Consultant (Note: – The Individual Consultant Agreements listed below are less than $5,000 per year)

Aptinyx, Inc., Atai Life Sciences, Biogen, Idec, MA, Bionomics, Limited (Australia), Boehringer Ingelheim International, Cadent Therapeutics, Inc., Clexio Bioscience, Ltd., COMPASS Pathways, Limited, United Kingdom, Concert Pharmaceuticals, Inc., Epiodyne, Inc., EpiVario, Inc., Greenwich Biosciences, Inc., Heptares Therapeutics, Limited (UK), Janssen Research & Development, Jazz Pharmaceuticals, Inc., Otsuka America Pharmaceutical, Inc., Perception Neuroscience Holdings, Inc., Spring Care, Inc., Sunovion Pharmaceuticals, Inc., Takeda Industries, Taisho Pharmaceutical Co., Ltd

### Board of Directors

Freedom Biosciences, Inc.

### Scientific Advisory Board

Biohaven Pharmaceuticals, BioXcel Therapeutics, Inc. (Clinical Advisory Board), Cadent Therapeutics, Inc. (Clinical Advisory Board), Cerevel Therapeutics, LLC, Delix Therapeutics, Inc., EpiVario, Inc., Eisai, Inc., Jazz Pharmaceuticals, Inc., Neurocrine Biosciences, Inc., Novartis Pharmaceuticals Corporation, PsychoGenics, Inc., Neumora Therapeutics, Inc., Tempero Bio, Inc., Terran Biosciences, Inc.

### Stock

Biohaven Pharmaceuticals, Sage Pharmaceuticals, Spring Care, Inc.

### Stock Options

Biohaven Pharmaceuticals Medical Sciences, EpiVario, Inc., Neumora Therapeutics, Inc., Terran Biosciences, Inc., Tempero Bio, Inc.

### Income Greater than $10,000

#### Editorial Board

Editor - Biological Psychiatry

### Patents and Inventions

1. Seibyl JP, Krystal JH, Charney DS. Dopamine and noradrenergic reuptake inhibitors in treatment of schizophrenia. US Patent #:5,447,948.September 5, 1995
2. Vladimir, Coric, Krystal, John H, Sanacora, Gerard – Glutamate Modulating Agents in the Treatment of Mental Disorders. US Patent No. 8,778,979 B2 Patent Issue Date: July 15, 2014. US Patent Application No. 15/695,164:
3. Filing Date: 09/05/2017
4. Charney D, Krystal JH, Manji H, Matthew S, Zarate C., - Intranasal Administration of Ketamine to Treat Depression United States Patent Number: 9592207, Issue date: 3/14/2017. Licensed to Janssen Research & Development
5. Zarate, C, Charney, DS, Manji, HK, Mathew, Sanjay J, Krystal, JH, Yale University “Methods for Treating Suicidal Ideation”, Patent Application No. 15/379,013 filed on December 14, 2016 by Yale University Office of Cooperative Research
6. Arias A, Petrakis I, Krystal JH. – Composition and methods to treat addiction.
7. Provisional Use Patent Application no.61/973/961. April 2, 2014. Filed by Yale University Office of Cooperative Research.
8. Chekroud, A., Gueorguieva, R., & Krystal, JH. “Treatment Selection for Major Depressive Disorder” [filing date 3^rd^ June 2016, USPTO docket number Y0087.70116US00]. Provisional patent submission by Yale University
9. Gihyun, Yoon, Petrakis I, Krystal JH – Compounds, Compositions and Methods for Treating or Preventing Depression and Other Diseases. U. S. Provisional Patent Application No. 62/444,552, filed on January10, 2017 by Yale University Office of Cooperative Research OCR 7088 US01
10. Abdallah, C, Krystal, JH, Duman, R, Sanacora, G. Combination Therapy for Treating or Preventing Depression or Other Mood Diseases. U.S. Provisional Patent Application No. 62/719,935 filed on August 20, 2018 by Yale University Office of Cooperative Research OCR 7451 US01
11. John Krystal, Godfrey Pearlson, Stephanie O’Malley, Marc Potenza, Fabrizio Gasparini, Baltazar Gomez-Mancilla, Vincent Malaterre. Mavoglurant in treating gambling and gaming disorders. U.S. Provisional Patent Application No. 63/125,181filed on December 14, 2020 by Yale University Office of Cooperative Research OCR 8065 US00

### NON Federal Research Support

AstraZeneca Pharmaceuticals provides the drug, Saracatinib, for research related to NIAAA grant “Center for Translational Neuroscience of Alcoholism [CTNA-4]

Novartis provides the drug, Mavoglurant, for research related to NIAAA grant “Center for Translational Neuroscience of Alcoholism [CTNA-4]

Cerevel provides the drug PF-06412562 for A Translational and Neurocomputational Evaluation of a D1R Partial Agonist for Schizophrenia (1 U01 MH121766-01)

## Notes

### Competing Interest Statement

The authors have declared no competing interest.

## REFERENCES

1. Widiger TA, Oltmanns JR (2017): Neuroticism is a fundamental domain of personality with enormous public health implications. World Psychiatry. 16:144–145.

2. Schmitt DP, Realo A, Voracek M, Allik J (2008): Why can’t a man be more like a woman? Sex differences in Big Five personality traits across 55 cultures. J Pers Soc Psychol. 94:168–182.

3. South SC, Jarnecke AM, Vize CE (2018): Sex differences in the Big Five model personality traits: A behavior genetics exploration. Journal of Research in Personality. 74:158–165.

4. Weisberg YJ, Deyoung CG, Hirsh JB (2011): Gender Differences in Personality across the Ten Aspects of the Big Five. Front Psychol. 2:178.

5. Hasin DS, Sarvet AL, Meyers JL, Saha TD, Ruan WJ, Stohl M, et al. (2018): Epidemiology of Adult DSM-5 Major Depressive Disorder and Its Specifiers in the United States. JAMA Psychiatry. 75:336–346.

6. Wendt FR, Pathak GA, Tylee DS, Goswami A, Polimanti R (2020): Heterogeneity and Polygenicity in Psychiatric Disorders: A Genome-Wide Perspective. Chronic Stress (Thousand Oaks). 4:2470547020924844.

7. Polimanti R, Wendt FR (2021): Posttraumatic stress disorder: from gene discovery to disease biology. Psychological Medicine. 51:2178–2188.

8. Vukasović T, Bratko D (2015): Heritability of personality: A meta-analysis of behavior genetic studies. Psychol Bull. 141:769–785.

9. Okbay A, Beauchamp JP, Fontana MA, Lee JJ, Pers TH, Rietveld CA, et al. (2016): Genome-wide association study identifies 74 loci associated with educational attainment. Nature. 533:539–542.

10. Power RA, Pluess M (2015): Heritability estimates of the Big Five personality traits based on common genetic variants. Transl Psychiatry. 5:e604.

11. Smith DJ, Escott-Price V, Davies G, Bailey ME, Colodro-Conde L, Ward J, et al. (2016): Genome-wide analysis of over 1061000 individuals identifies 9 neuroticism-associated loci. Mol Psychiatry. 21:749–757.

12. Vinkhuyzen AA, Pedersen NL, Yang J, Lee SH, Magnusson PK, Iacono WG, et al. (2012): Common SNPs explain some of the variation in the personality dimensions of neuroticism and extraversion. Transl Psychiatry. 2:e102.

13. Luciano M, Davies G, Summers KM, Hill WD, Hayward C, Liewald DC, et al. (2021): The influence of X chromosome variants on trait neuroticism. Molecular Psychiatry. 26:483–491.

14. Nagel M, Jansen PR, Stringer S, Watanabe K, de Leeuw CA, Bryois J, et al. (2018): Meta-analysis of genome-wide association studies for neuroticism in 449,484 individuals identifies novel genetic loci and pathways. Nat Genet. 50:920–927.

15. Turley P, Walters RK, Maghzian O, Okbay A, Lee JJ, Fontana MA, et al. (2018): Multi-trait analysis of genome-wide association summary statistics using MTAG. Nat Genet. 50:229–237.

16. Wendt FR, Pathak GA, Lencz T, Krystal JH, Gelernter J, Polimanti R (2021): Multivariate genome-wide analysis of education, socioeconomic status and brain phenome. Nature Human Behaviour. 5:482– 496.

17. Hill WD, Weiss A, Liewald DC, Davies G, Porteous DJ, Hayward C, et al. (2019): Genetic contributions to two special factors of neuroticism are associated with affluence, higher intelligence, better health, and longer life. Mol Psychiatry.

18. Eysenck HJ, Eysenck SBG (1975): Manual of the Eysenck Personality Questionnaire. London: Hodder and Stoughton.

19. Smith DJ, Nicholl BI, Cullen B, Martin D, Ul-Haq Z, Evans J, et al. (2013): Prevalence and Characteristics of Probable Major Depression and Bipolar Disorder within UK Biobank: Cross-Sectional Study of 172,751 Participants. PLOS ONE. 8:e75362.

20. Bycroft C, Freeman C, Petkova D, Band G, Elliott LT, Sharp K, et al. (2018): The UK Biobank resource with deep phenotyping and genomic data. Nature. 562:203–209.

21. Chang CC, Chow CC, Tellier LC, Vattikuti S, Purcell SM, Lee JJ (2015): Second-generation PLINK: rising to the challenge of larger and richer datasets. Gigascience. 4:7.

22. Gao F, Chang D, Biddanda A, Ma L, Guo Y, Zhou Z, et al. (2015): XWAS: A Software Toolset for Genetic Data Analysis and Association Studies of the X Chromosome. Journal of Heredity. 106:666–671.

23. Purcell S, Neale B, Todd-Brown K, Thomas L, Ferreira MA, Bender D, et al. (2007): PLINK: a tool set for whole-genome association and population-based linkage analyses. Am J Hum Genet. 81:559–575.

24. Choi SW, O’Reilly PF (2019): PRSice-2: Polygenic Risk Score software for biobank-scale data. Gigascience. 8.

25. Bulik-Sullivan BK, Loh PR, Finucane HK, Ripke S, Yang J, Patterson N, et al. (2015): LD Score regression distinguishes confounding from polygenicity in genome-wide association studies. Nat Genet. 47:291–295.

26. Finucane HK, Bulik-Sullivan B, Gusev A, Trynka G, Reshef Y, Loh PR, et al. (2015): Partitioning heritability by functional annotation using genome-wide association summary statistics. Nat Genet. 47:1228–1235.

27. Gazal S, Marquez-Luna C, Finucane HK, Price AL (2019): Reconciling S-LDSC and LDAK functional enrichment estimates. Nat Genet. 51:1202–1204.

28. Wendt FR, Pathak GA, Overstreet C, Tylee DS, Gelernter J, Atkinson EG, et al. (2021): Characterizing the effect of background selection on the polygenicity of brain-related traits. Genomics. 113:111–119.

29. Watanabe K, Taskesen E, van Bochoven A, Posthuma D (2017): Functional mapping and annotation of genetic associations with FUMA. Nat Commun. 8:1826.

30. Watanabe K, Umicevic Mirkov M, de Leeuw CA, van den Heuvel MP, Posthuma D (2019): Genetic mapping of cell type specificity for complex traits. Nat Commun. 10:3222.

31. Ge T, Chen CY, Ni Y, Feng YA, Smoller JW (2019): Polygenic prediction via Bayesian regression and continuous shrinkage priors. Nat Commun. 10:1776.

32. Dennis JK, Sealock JM, Straub P, Lee YH, Hucks D, Actkins KE, et al. (2021): Clinical laboratory test-wide association scan of polygenic scores identifies biomarkers of complex disease. Genome Medicine. 13:6.

33. Roden DM, Pulley JM, Basford MA, Bernard GR, Clayton EW, Balser JR, et al. (2008): Development of a large-scale de-identified DNA biobank to enable personalized medicine. Clin Pharmacol Ther. 84:362–369.

34. Wu P, Gifford A, Meng X, Li X, Campbell H, Varley T, et al. (2019): Mapping ICD-10 and ICD-10-CM Codes to Phecodes: Workflow Development and Initial Evaluation. JMIR Med Inform. 7:e14325.

35. Gusev A, Ko A, Shi H, Bhatia G, Chung W, Penninx BW, et al. (2016): Integrative approaches for large-scale transcriptome-wide association studies. Nat Genet. 48:245–252.

36. Kang HJ, Kawasawa YI, Cheng F, Zhu Y, Xu X, Li M, et al. (2011): Spatio-temporal transcriptome of the human brain. Nature. 478:483–489.

37. Lee JJ, Wedow R, Okbay A, Kong E, Maghzian O, Zacher M, et al. (2018): Gene discovery and polygenic prediction from a genome-wide association study of educational attainment in 1.1 million individuals. Nat Genet. 50:1112–1121.

38. Gelernter J, Sun N, Polimanti R, Pietrzak R, Levey DF, Bryois J, et al. (2019): Genome-wide association study of post-traumatic stress disorder reexperiencing symptoms in >165,000 US veterans. Nat Neurosci. 22:1394–1401.

39. Stein MB, Levey DF, Cheng Z, Wendt FR, Harrington K, Pathak GA, et al. (2021): Genome-wide association analyses of post-traumatic stress disorder and its symptom subdomains in the Million Veteran Program. Nature Genetics. 53:174–184.

40. Power RA, Pluess M (2015): Heritability estimates of the Big Five personality traits based on common genetic variants. Translational Psychiatry. 5:e604–e604.

41. Tørring PM, Larsen MJ, Brasch-Andersen C, Krogh LN, Kibæk M, Laulund L, et al. (2019): Is MED13L-related intellectual disability a recognizable syndrome? Eur J Med Genet. 62:129–136.

42. Strawbridge RJ, Ward J, Ferguson A, Graham N, Shaw RJ, Cullen B, et al. (2019): Identification of novel genome-wide associations for suicidality in UK Biobank, genetic correlation with psychiatric disorders and polygenic association with completed suicide. EBioMedicine. 41:517–525.

43. Wendt FR, Pathak GA, Levey DF, Nuñez YZ, Overstreet C, Tyrrell C, et al. (2021): Sex-stratified gene-by-environment genome-wide interaction study of trauma, posttraumatic-stress, and suicidality. Neurobiol Stress. 14:100309.

44. Deneault E, Faheem M, White SH, Rodrigues DC, Sun S, Wei W, et al. (2019): CNTN5(-)(/+)or EHMT2(-)(/+)human iPSC-derived neurons from individuals with autism develop hyperactive neuronal networks. Elife. 8.

45. Camacho-Arroyo I, Piña-Medina AG, Bello-Alvarez C, Zamora-Sánchez CJ (2020): Chapter Seven - Sex hormones and proteins involved in brain plasticity. In: Litwack G, editor. Vitamins and Hormones: Academic Press, pp 145–165.

46. Edwards AC, Aliev F, Bierut LJ, Bucholz KK, Edenberg H, Hesselbrock V, et al. (2012): Genome-wide association study of comorbid depressive syndrome and alcohol dependence. Psychiatr Genet. 22:31–41.

47. Privé F, Aschard H, Carmi S, Folkersen L, Hoggart C, O’Reilly PF, et al. (2022): Portability of 245 polygenic scores when derived from the UK Biobank and applied to 9 ancestry groups from the same cohort. The American Journal of Human Genetics. 109:12–23.

48. Sirugo G, Williams SM, Tishkoff SA (2019): The Missing Diversity in Human Genetic Studies. Cell. 177:26–31.

49. Vasdev N, Kumar A, Veeratterapillay R, Thorpe AC (2013): Hematuria secondary to benign prostatic hyperplasia: retrospective analysis of 166 men identified in a single one stop hematuria clinic. Curr Urol. 6:146–149.

50. Perry LM, Hoerger M, Silberstein J, Sartor O, Duberstein P (2018): Understanding the distressed prostate cancer patient: Role of personality. Psychooncology. 27:810–816.

51. Weuve J, Mendes de Leon CF, Bennett DA, Dong X, Evans DA (2014): The red cell distribution width and anemia in association with prevalent dementia. Alzheimer Dis Assoc Disord. 28:99–105.

52. Yamaguchi S, Hamano T, Oka T, Doi Y, Kajimoto S, Shimada K, et al. (2021): Mean corpuscular hemoglobin concentration: an anemia parameter predicting cardiovascular disease in incident dialysis patients. J Nephrol.

53. Just MJ, Kozakiewicz M (2015): Depressive disorders co-existing with Addison-Biermer anemia - case report. Neuropsychiatr Dis Treat. 11:1145–1148.

54. Bahrami A, Khorasanchi Z, Tayefi M, Avan A, Seifi N, Tavakoly Sany SB, et al. (2020): Anemia is associated with cognitive impairment in adolescent girls: A cross-sectional survey. Applied Neuropsychology: Child. 9:165–171.

55. Lee H-S, Chao H-H, Huang W-T, Chen SC-C, Yang H-Y (2020): Psychiatric disorders risk in patients with iron deficiency anemia and association with iron supplementation medications: a nationwide database analysis. BMC Psychiatry. 20:216.

56. Sobrero A, Puglisi F, Guglielmi A, Belvedere O, Aprile G, Ramello M, et al. (2001): Fatigue: a main component of anemia symptomatology. Semin Oncol. 28:15–18.

57. Hidese S, Saito K, Asano S, Kunugi H (2018): Association between iron-deficiency anemia and depression: A web-based Japanese investigation. Psychiatry and Clinical Neurosciences. 72:513–521.

58. Girgenti MJ, Wang J, Ji D, Cruz DA, Stein MB, Gelernter J, et al. (2021): Transcriptomic organization of the human brain in post-traumatic stress disorder. Nat Neurosci. 24:24–33.

59. Levey DF, Stein MB, Wendt FR, Pathak GA, Zhou H, Aslan M, et al. (2021): Bi-ancestral depression GWAS in the Million Veteran Program and meta-analysis in >1.2 million individuals highlight new therapeutic directions. Nat Neurosci. 24:954–963.

60. Gaspar HA, Gerring Z, Hübel C, Middeldorp CM, Derks EM, Breen G, et al. (2019): Using genetic drug-target networks to develop new drug hypotheses for major depressive disorder. Translational Psychiatry. 9:117.

61. Kuwahara T, Inoue K, D’Agati VD, Fujimoto T, Eguchi T, Saha S, et al. (2016): LRRK2 and RAB7L1 coordinately regulate axonal morphology and lysosome integrity in diverse cellular contexts. Scientific Reports. 6:29945.

62. Hindley G, Bahrami S, Steen NE, O’Connell KS, Frei O, Shadrin A, et al. (2021): Characterising the shared genetic determinants of bipolar disorder, schizophrenia and risk-taking. Transl Psychiatry. 11:466.

63. MacLeod DA, Rhinn H, Kuwahara T, Zolin A, Di Paolo G, McCabe BD, et al. (2013): RAB7L1 interacts with LRRK2 to modify intraneuronal protein sorting and Parkinson’s disease risk. Neuron. 77:425–439.

64. Strom NI, Yu D, Gerring ZF, Halvorsen MW, Abdellaoui A, Rodriguez-Fontenla C, et al. (2021): Genome-wide association study identifies new locus associated with OCD. medRxiv.2021.2010.2013.21261078.

65. Zhong J, Li S, Zeng W, Li X, Gu C, Liu J, et al. (2019): Integration of GWAS and brain eQTL identifies FLOT1 as a risk gene for major depressive disorder. Neuropsychopharmacology. 44:1542–1551.

66. Reisinger SN, Kong E, Molz B, Humberg T, Sideromenos S, Cicvaric A, et al. (2019): Flotillin-1 interacts with the serotonin transporter and modulates chronic corticosterone response. Genes Brain Behav. 18:e12482.

67. Montoliu T, Hidalgo V, Salvador A (2020): Personality and Hypothalamic–Pituitary–Adrenal Axis in Older Men and Women. Frontiers in Psychology. 11.

68. Qin D-d, Rizak J, Feng X-l, Yang S-c, Lü L-b, Pan L, et al. (2016): Prolonged secretion of cortisol as a possible mechanism underlying stress and depressive behaviour. Scientific Reports. 6:30187.

69. Powers SI, Laurent HK, Gunlicks-Stoessel M, Balaban S, Bent E (2016): Depression and anxiety predict sex-specific cortisol responses to interpersonal stress. Psychoneuroendocrinology. 69:172–179.

