## Supplementary Material for "Sex-specific genetic and transcriptomic liability to neuroticism"

Short title: sex-specific multi-omic liability to neuroticism

Keywords: neuroticism, sex-effects, GWAS, TWAS, pituitary gland, biomarkers

### **SUPPLMENTARY INFORMATION**

**METHODS**

**Linkage Disequilibrium Score Regression (LDSC) – Partitioned Heritability**

To test which functional categories contribute to the *h^2^* of neuroticism, we used LDSC to partition each *h^2^* estimate using 118 genomic annotations. Briefly, partitioned *h^2^* assumes a linear model of trait *h^2^* where the *h^2^* per genomic annotation is the sum of squared effect sizes per SNP belonging to each genomic annotation (1). The 118 annotations used in this study are derived from baseline annotations and expanded genomic annotations. Baseline and expanded annotation include coding, untranslated, promoter, and intronic regions, histone methylation and acetylation sites, chromatin states including DNase-I hypersensitivity regions, regions conserved in mammals, super-enhancers, allele frequency strata, etc. (1, 2). We included additional annotations for loss-of-function intolerance, positively selected, negatively selected, and Neanderthal introgressed regions. All genomic annotations for partitioned *h^2^* have been described previously (1-3). Because of the nature of the test, p-values derived from partitioned *h^2^* linear models were not subjected to multiple testing correction.

**REFERENCES**

1.	Finucane HK, Bulik-Sullivan B, Gusev A, Trynka G, Reshef Y, Loh PR, et al. (2015): Partitioning heritability by functional annotation using genome-wide association summary statistics. *Nat Genet*. 47:1228-1235.

2.	Gazal S, Marquez-Luna C, Finucane HK, Price AL (2019): Reconciling S-LDSC and LDAK functional enrichment estimates. *Nat Genet*. 51:1202-1204.

3.	Wendt FR, Pathak GA, Overstreet C, Tylee DS, Gelernter J, Atkinson EG, et al. (2021): Characterizing the effect of background selection on the polygenicity of brain-related traits. *Genomics*. 113:111-119.

**FIGURES**

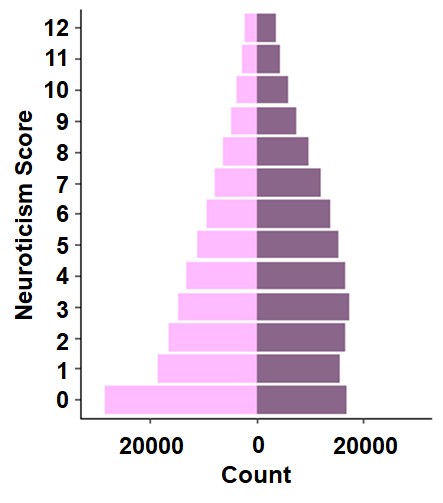

**Fig S1.** Distribution of neuroticism scores in UK Biobank males (right) and females (left) of European descent.

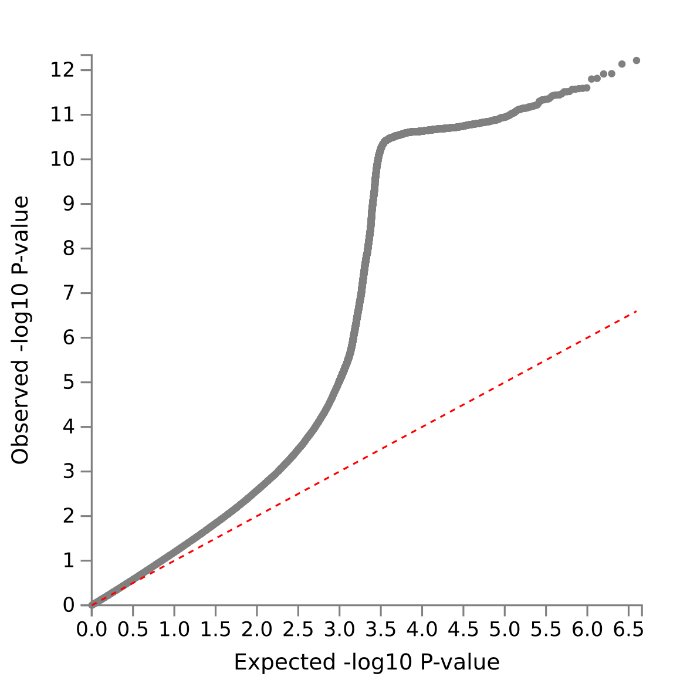

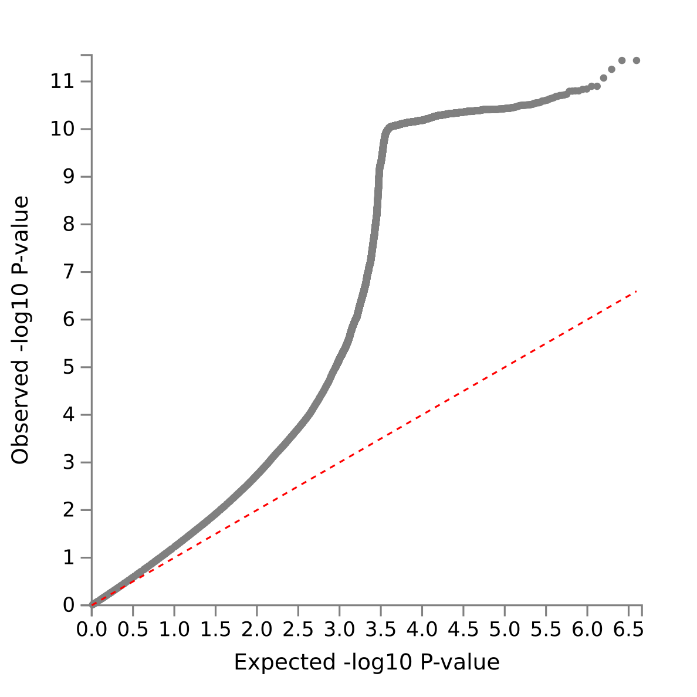

**Fig S2. Quantile-quantile plots.** QQ plots of male (left) and female (right) neuroticism GWAS in the UK Biobank.

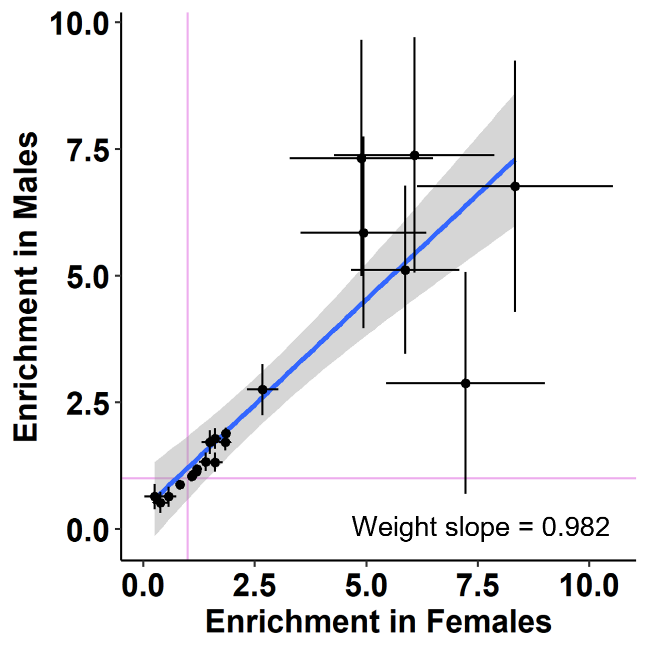

**Fig S3. Partitioned *h^2^* comparison.** Overlapping enrichment of nominally significant genomic annotations in male and female GWAS of neuroticism. Each data point is a genomic annotation with the standard error shown as horizontal (female) and vertical (male) bars. Purple lines indicate intercepts of 1 which are interpreted as no enrichment. The dashed grey line denotes the 1-to-1 relationship between males and females and the blue line shows the linear regression between sexes.

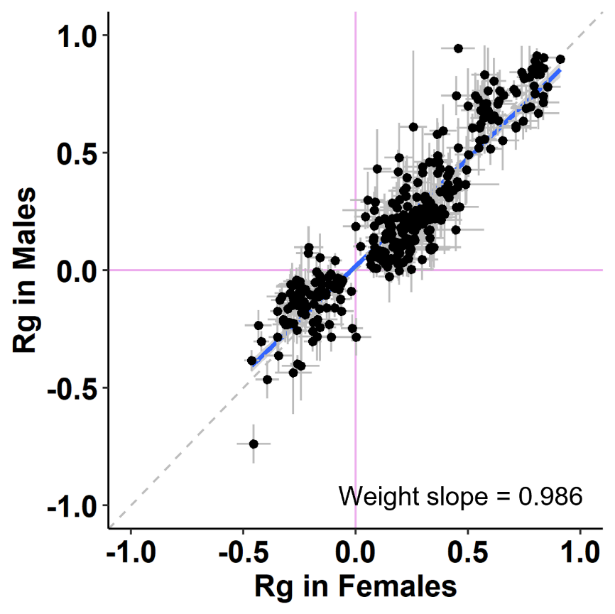

**Fig S4. Genetic correlation comparison.** Relationship between males and females for all nominally significant genetic correlations (r_g_). Each data point is the r_g_ between neuroticism and a trait from the UK Biobank with the standard error shown as horizontal (female) and vertical (male) bars. Purple lines indicate r_g_ of 0. The dashed grey line denotes the 1-to-1 relationship between males and females and the blue line shows the linear regression between sexes.

**TABLES**

**Table S1.** Genome-wide significant loci associated with neuroticism in males and females. Highlighted rows show the six loci meeting genome-wide significance (*P* < 5x10^-8^) in one sex and failing to meet nominal significance in the other sex (*P* > 0.05); for these loci, two-sided Z-test results are shown to demonstrate the sex-specificity of their effects.

**Table S2.** Positional mapping results for male and female neuroticism risk loci.

**Table S3.** Cross-ancestry polygenic scoring (PGS) results. The highlighted row shows the analysis significant in one sex but not in the other (*P* > 0.05); for this result, a two-sided Z-test was performed to test the sex-specificity of these PGS association.

**Table S4.** Partitioned heritability results for neuroticism in males and females. Highlighted rows show the genomic partitions enriched in one sex but not in the other (*P* > 0.05); for these genomic annotations, two-sided Z-test results are shown to test the sex-specificity of these enrichments.

**Table S5.** Genetic correlation between UK Biobank phenotypes and neuroticism tested in males and females. Highlighted rows show the significant genetic correlations in one sex but not in the other (*P* > 0.05); for these results, two-sided Z-test results are shown to test the sex-specificity of these genetic correlations.

**Table S6**. Phenome-wide association of neuroticism polygenic scores and phecodes from the Vanderbilt University Biobank. Highlighted rows show the significant associations in one sex but not in the other (*P* > 0.05); for these results, two-sided Z-test results are shown to test the sex-specificity of these associations.

**Table S7**. Laboratory-measure-wide association of neuroticism polygenic scores and phecodes from the Vanderbilt University Biobank. Highlighted rows show the significant associations in one sex but not in the other (*P* > 0.05); for these results, two-sided Z-test results are shown to test the sex-specificity of these associations.

**Table S8**. Enrichment of tissue transcriptomic profiles in males and females. Highlighted rows show the significant tissue transcriptomic enrichments in one sex but not in the other (*P* > 0.05); for these results, two-sided Z-test results are shown to test the sex-specificity of these enrichments.

**Table S9**. Transcriptome-wide association study of neuroticism in the frontal cortex (BA9) of GTEx v8. Highlighted rows show the genes associated with neuroticism in one sex but not in the other (*P* > 0.05).

**Table S10**. (A) BrainSpan results for frontal cortex gene expression at ZNF204P, HTR1D, and DCAKD across developmental stages. (B) Mean expression of HTR1D across developmental stage and sex.

**Table S11**. Transcriptome-wide association study of neuroticism in the pituitary gland of GTEx v8. Highlighted rows show the genes associated with neuroticism in one sex but not in the other (*P* > 0.05).
